# Transmission dynamics of COVID-19 in Ghana and the impact of public health interventions

**DOI:** 10.1101/2021.07.04.21259991

**Authors:** Sylvia K. Ofori, Jessica S. Schwind, Kelly L. Sullivan, Benjamin J. Cowling, Gerardo Chowell, Isaac Chun-Hai Fung

## Abstract

The study characterized COVID-19 transmission in Ghana in 2020-21 by estimating the time-varying reproduction number (R_t_) and exploring its association with various public health interventions at the national and regional levels. Ghana experienced four pandemic waves with epidemic peaks in July 2020, and January, August and December, 2021. The epidemic peak was the highest nationwide in December 2021 with R_t_ ≥2. Throughout 2020-21, per-capita cumulative case count by region increased with population size. Mobility data suggested negative correlation between R_t_ and staying home in the first 90 days of the pandemic. The relaxation of movement restrictions and religious gatherings were not associated with increased R_t_ in the regions with lower case burdens. R_t_ decreased from above 1 when schools reopened in January 2021 to below 1 after vaccination rollout in March 2021. Findings indicated most public health interventions were associated with R_t_ reduction at the national and regional levels.

## Main text

As of January 15, 2022, 153,514 cases of COVID-19 were confirmed in Ghana, with 1,343 deaths and 9,020 active cases.^1^ Among African countries, Ghana has controlled COVID-19 transmission with a substantial package of public health and social measures (Oxford stringency index of 62.04 as of May 7, 2020), ^2,3^ including a 14-day mandatory quarantine for all persons who entered the country, school and church closures, a lockdown of major cities, and internal movement restrictions.^4^

This study described the COVID-19 transmission dynamics in Ghana by estimating the time-varying reproduction number (R_t_) using Cori et al method^5^ and Generalized Growth Model (GGM) (Supplementary Materials)^6^ to assess the impact of interventions at the national and regional levels and to explore the association between population size and the COVID-19 cumulative incidence. The correlation between population mobility and incident case count, and between mobility and R_t_ was also assessed.

The daily number of new infections and daily cumulative incidence data by date of report for Ghana and each of its sixteen regions was obtained through the Johns Hopkins University COVID-19 dashboard from March 12, 2020, to December 31, 2021 (Table S1, Figures S1 and S2). To account for testing delay (three days) and incubation period (six days), the time series was shifted by nine days to approximate the date of infection.^8^ Using three-day moving averages of interpolated daily incident case count data, we used the EpiEstim package in R version 4.0.3 to estimate the R_t_ using the 7-day sliding window and the nonoverlapping time window between interventions.^5^ To compare the R_t_ before, during, and after policies were implemented and assess their association with COVID-19 transmission, specific time points at which a bundle of interventions began were selected for the latter analysis and the average R_t_ estimates over the period between two policy change time points were estimated (Table 1).

**Table 1:**
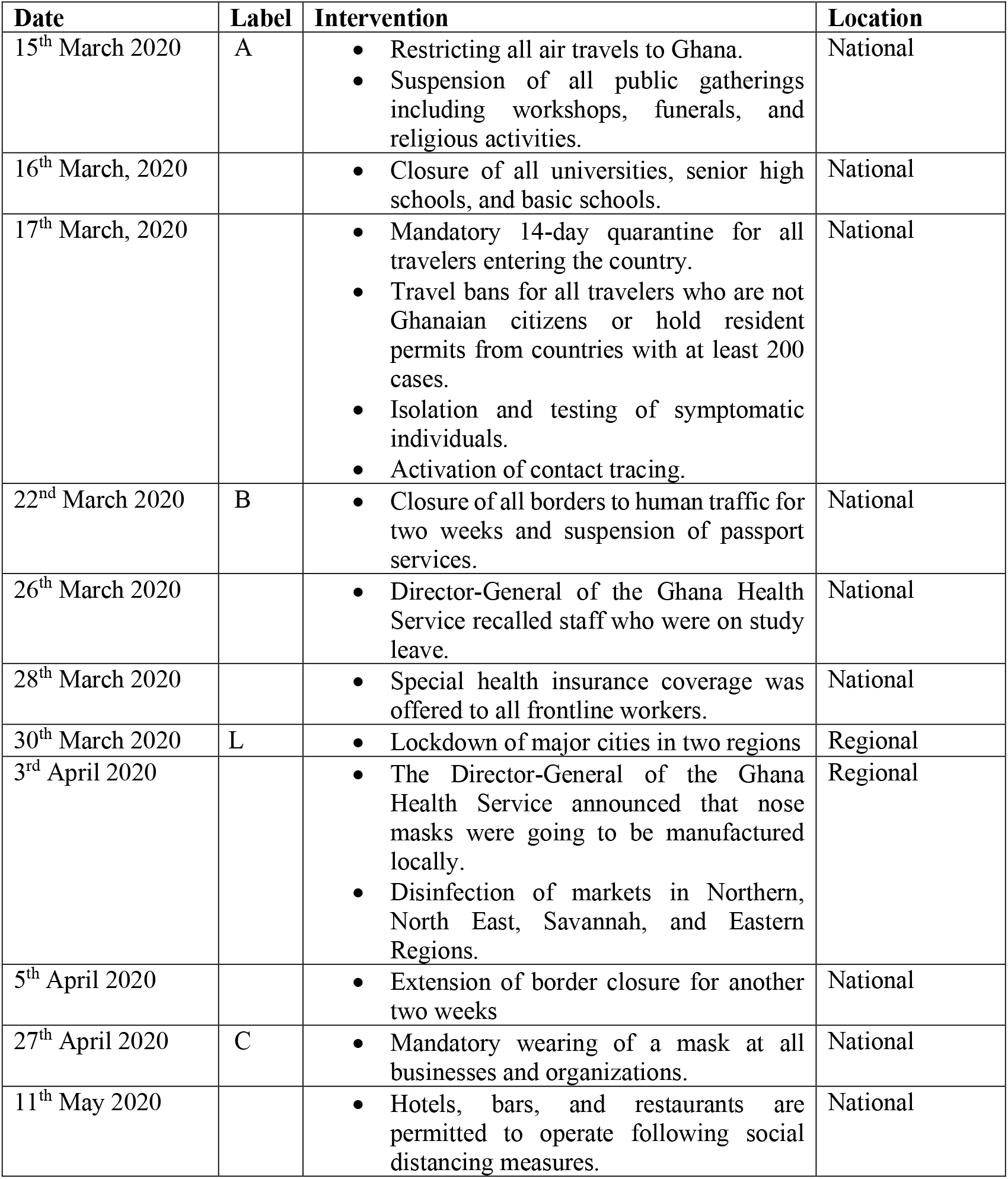
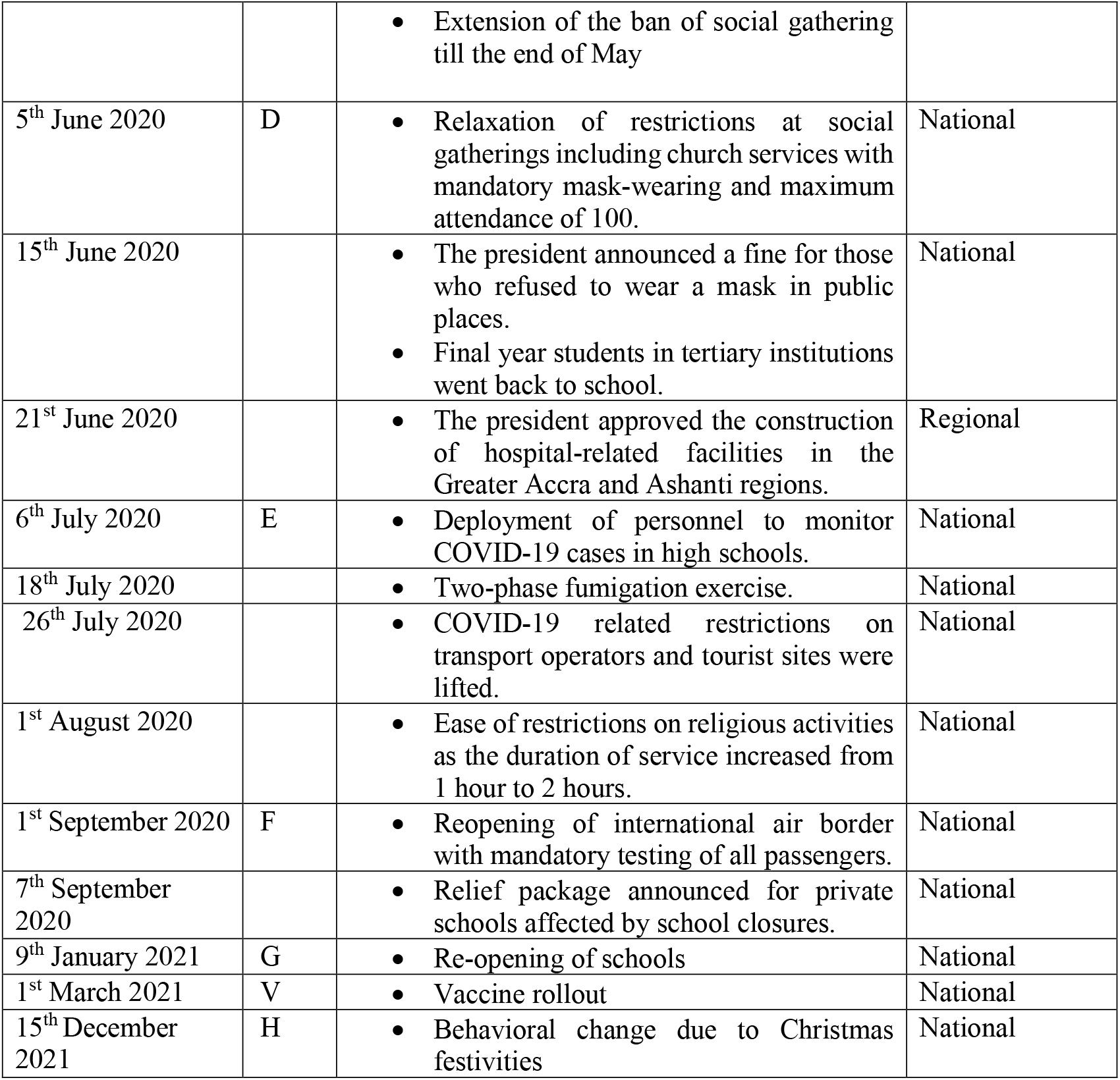
Public health and social measures interventions against COVID-19 implemented in Ghana^1,21–23^.

| **Date** | **Label** | **Intervention** | **Location** |
| --- | --- | --- | --- |
| 15<sup>th</sup> March 2020 | A | Restricting all air travels to Ghana.<br>Suspension of all public gatherings including workshops, funerals, and religious activities. | National |
| 16<sup>th</sup> March, 2020 |  | Closure of all universities, senior high schools, and basic schools. | National |
| 17<sup>th</sup> March, 2020 |  | Mandatory 14-day quarantine for all travelers entering the country.<br>Travel bans for all travelers who are not Ghanaian citizens or hold resident permits from countries with at least 200 cases.<br>Isolation and testing of symptomatic individuals.<br>Activation of contact tracing. | National |
| 22<sup>nd</sup> March 2020 | B | Closure of all borders to human traffic for two weeks and suspension of passport services. | National |
| 26<sup>th</sup> March 2020 |  | Director-General of the Ghana Health Service recalled staff who were on study leave. | National |
| 28<sup>th</sup> March 2020 |  | Special health insurance coverage was offered to all frontline workers. | National |
| 30<sup>th</sup> March 2020 | L | Lockdown of major cities in two regions | Regional |
| 3<sup>rd</sup> April 2020 |  | The Director-General of the Ghana Health Service announced that nose masks were going to be manufactured locally.<br>Disinfection of markets in Northern, North East, Savannah, and Eastern Regions. | Regional |
| 5<sup>th</sup> April 2020 |  | Extension of border closure for another two weeks | National |
| 27<sup>th</sup> April 2020 | C | Mandatory wearing of a mask at all businesses and organizations. | National |
| 11<sup>th</sup> May 2020 |  | Hotels, bars, and restaurants are permitted to operate following social distancing measures. | National |
|  |  | Extension of the ban of social gathering till the end of May |  |
| 5<sup>th</sup> June 2020 | D | Relaxation of restrictions at social gatherings including church services with mandatory mask-wearing and maximum attendance of 100. | National |
| 15<sup>th</sup> June 2020 |  | The president announced a fine for those who refused to wear a mask in public places.<br>Final year students in tertiary institutions went back to school. | National |
| 21<sup>st</sup> June 2020 |  | The president approved the construction of hospital-related facilities in the Greater Accra and Ashanti regions. | Regional |
| 6<sup>th</sup> July 2020 | E | Deployment of personnel to monitor COVID-19 cases in high schools. | National |
| 18<sup>th</sup> July 2020 |  | Two-phase fumigation exercise. | National |
| 26<sup>th</sup> July 2020 |  | COVID-19 related restrictions on transport operators and tourist sites were lifted. | National |
| 1<sup>st</sup> August 2020 |  | Ease of restrictions on religious activities as the duration of service increased from 1 hour to 2 hours. | National |
| 1<sup>st</sup> September 2020 | F | Reopening of international air border with mandatory testing of all passengers. | National |
| 7<sup>th</sup> September 2020 |  | Relief package announced for private schools affected by school closures. | National |
| 9<sup>th</sup> January 2021 | G | Re-opening of schools | National |
| 1<sup>st</sup> March 2021 | V | Vaccine rollout | National |
| 15<sup>th</sup> December 2021 | H | Behavioral change due to Christmas festivities | National |

The power-law relationship between cumulative case number and population size was investigated at ten time points in 2020-21, using linear regression of log_10_-transformed per-capita cumulative case number and log_10_-transformed population size.^9^ The relationship between 7-day moving average of mobility changes and the 3-day moving average of daily number of new infections and that between 7-day moving average of mobility changes and the 7-day sliding window R_t_ in the first 90 days of the pandemic were assessed using the time-lagged cross-correlation with data from Google Mobility Report.^10^ The Georgia Southern University Institutional Review Board determined a non-human subject status for this project (H20364) under the G8 exemption category.

Ghana and its regions experienced four pandemic waves in 2020-21 with epidemic peaks in July 2020, and January, August and December, 2021. At the national level and in the Greater Accra and Ashanti regions, the R_t_ fluctuated around 1, increased to >1 before the epidemic peaks, and dropped <1 afterwards (Figure 1). The epidemic peak was the highest nationwide and in Greater Accra region in December 2021 with an estimated R_t_ of ≥2. Ghana’s December 2021 epidemic peak was largely driven by the Greater Accra region, while in other regions, case counts were increasing fast. Similar patterns in epidemic curves and R_t_ estimates were observed in Central, Eastern, Volta and Western regions, with R_t_ ≥3 in December 2021 (Figure S3).

**Figure 1:**
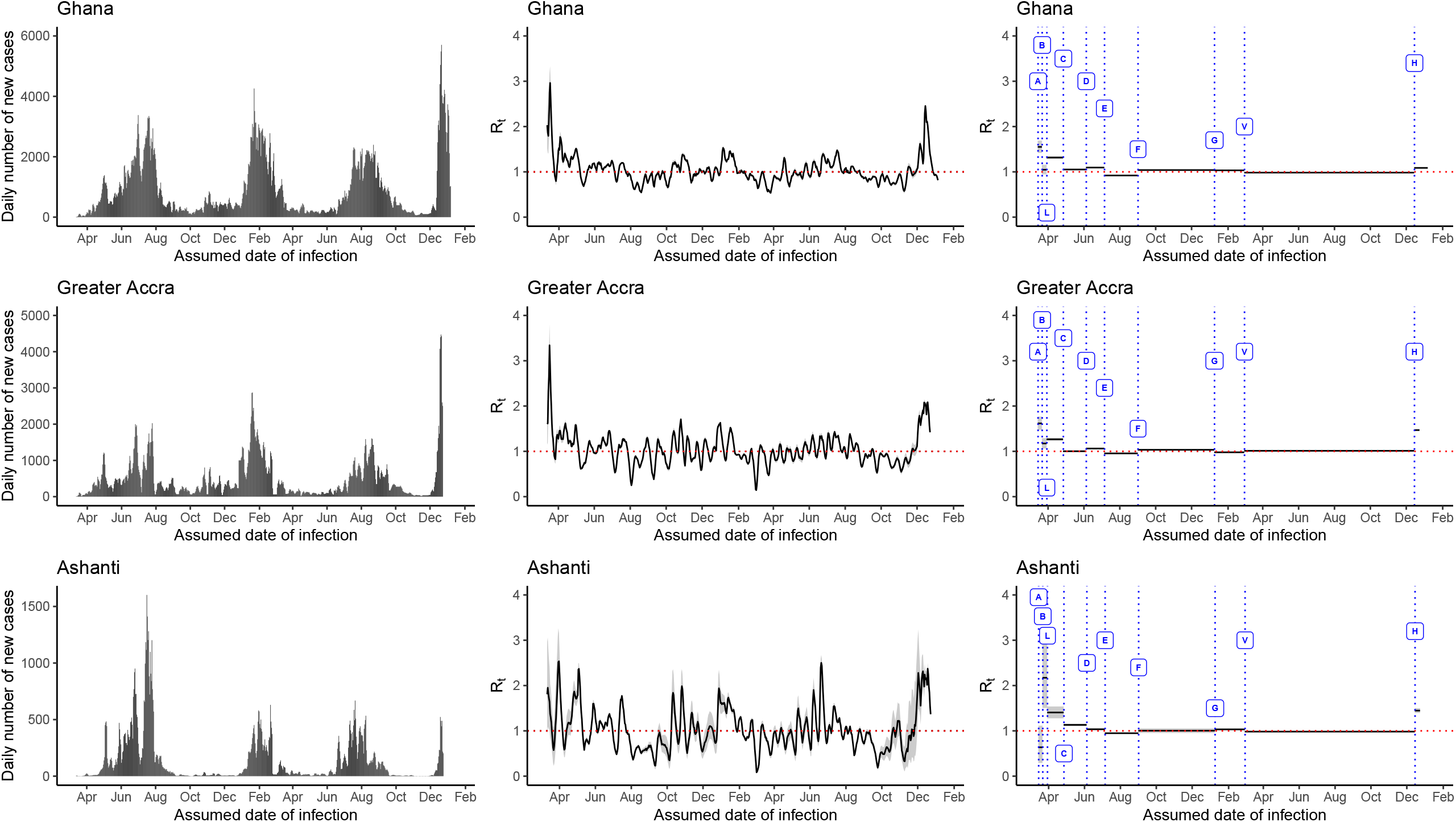
The daily number of new cases (left panels), 7-day sliding window *R*_*t*_ (middle panels) and nonoverlapping window *R*_*t*_ (right panels) estimated using the in the ‘EpiEstim’ package in Ghana, Greater Accra, and the Ashanti region, March 12, 2020—December 31, 2021. The government policies represented by the alphabets in the figure are: A = restrictions of all air travels to Ghana, suspension of social gatherings, school closure, mandatory 14-day quarantine for all travelers, B = closure of all borders to human traffics, L= lockdown of major cities, C = mandatory wearing of a mask at all businesses and organizations, D = relaxation of restrictions at social gatherings, E = deployment of personnel to monitor COVID-19 cases in high schools, F = reopening of international borders, G = Re-opening of schools, V = vaccination rollout, H= Christmas festivities in 2021. *Greater Accra and Ashanti regions are highlighted because they are the most populous and had the highest case burden in Ghana

The R_t_ estimates obtained using nonoverlapping windows suggested varying associations between government policies and interventions and the increase and decrease of COVID-19 transmission in Ghana and across regions. The restriction of social gatherings and travel bans implemented on March 15th, 2020, were associated with insignificant changes in R_t_ at the national level and the Greater Accra region. However, when all borders were closed to human traffic nationwide a week later, the R_t_ lowered to around 1 accounting for a 32.63% (95% credible interval, CrI: 22.87%, 41.19%) decrease for Ghana and a 27.41% (95% CrI: 16.68%, 36.26%) decrease in Greater Accra region. In contrast, the R_t_ for Ashanti increased by over 100%. On April 27th, 2020, the mandatory wearing of masks at all businesses and organizations was implemented nationwide which was associated with a decrease in R_t_ by 19.97% (95% CrI: 18.05%, 21.81%) at the national level. Ashanti and Greater Accra regions also observed significant declines in R_t_. The relaxation of restrictions on social gatherings was associated with a slight increase in R_t_ by 4.03% (95% CrI: 2.61%, 5.49%) for Ghana at the national level and by 6.02% (95% CrI: 4.05%, 7.78%) in the Greater Accra region, however the other regions observed a decline in R_t_ by >5%. Reopening of schools in January 2021 was associated with an increase in transmission in Eastern and Central regions only. Vaccination rollout was associated with a decline in R_t_ to <1 in Ghana and the regions except Greater Accra region which observed about a 3% increase. Overall, the behavioral change due to Christmas festivities in 2021 was associated with sustained transmission in all regions. Details of percent changes in R_t_ are in Table S2.

The assessment of population size and cumulative incidence showed that Ghanaian regions with larger populations experienced higher COVID-19 attack rates (Figure 2 and Table S3). Furthermore, there was a weak correlation between mobility changes and COVID-19 incidence in the first 90 days of the pandemic in Ghana (Tables S4 and S5), e.g., the daily number of new infections was positively correlated with mobility changes to retails and recreation, grocery and pharmacy and workplaces 3 days later (r>0, p<0.05 in all 3 cases), but no significant correlation was observed with residential mobility changes. R_t_ was negatively correlated with mobility changes to residence (r=−0.328, p=0.002).

**Figure 2:**
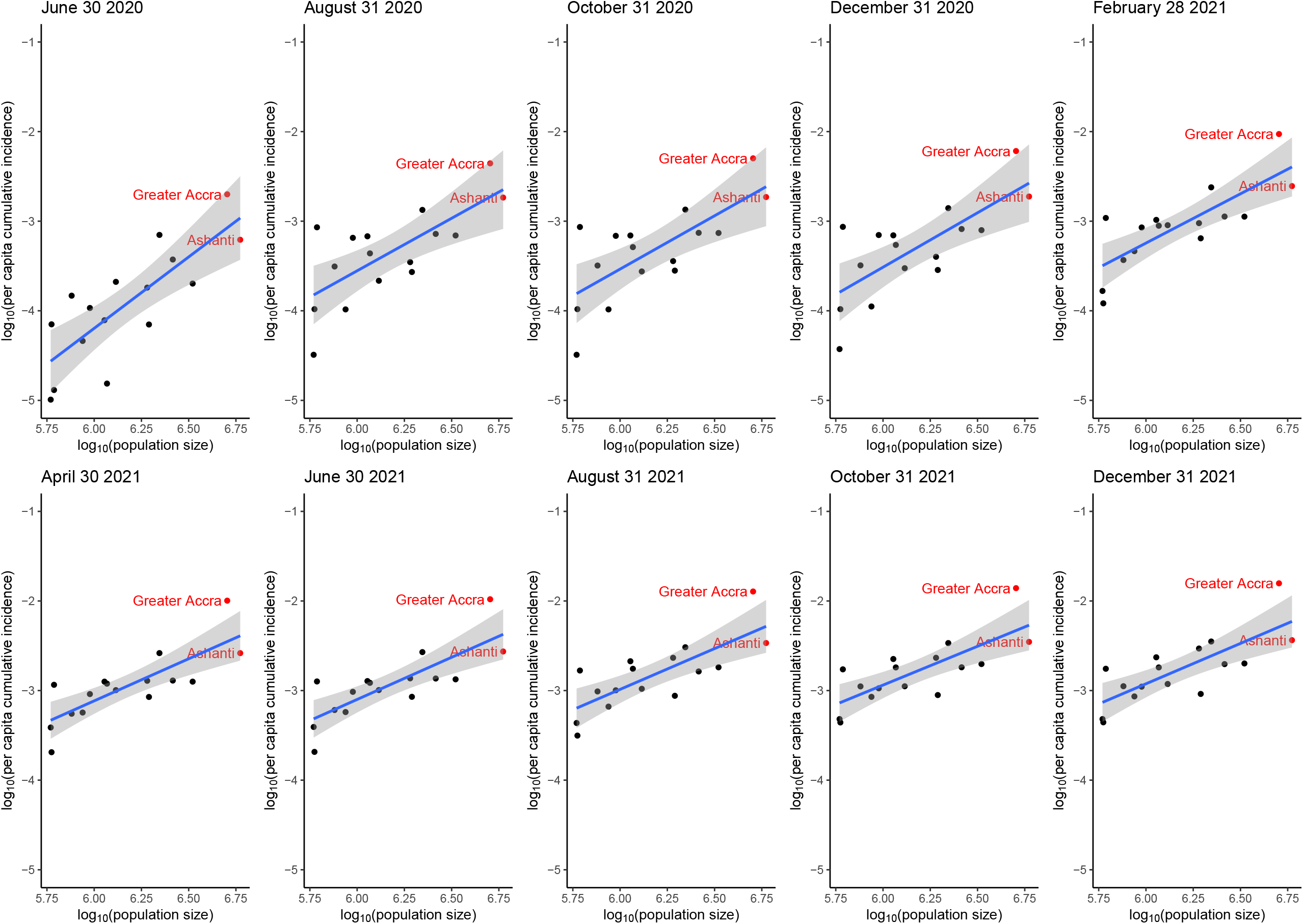
Linear regression models between log_10_-transformed per capita cumulative case count and log_10_-transformed population size of the sixteen regions of Ghana (grey areas representing the 95% confidence intervals of the regression lines) at 10-time points: June 30, August 31, October 31, December 31, 2020, and February 28, April 30, June 30, August 31, October 31, and December 31, 2021.

Overall, R_t_ >1 showed sustained transmission across the country, but some interventions implemented during the study period were associated with reduced R_t_ in regions with relatively higher case counts. Interestingly, relaxation of restrictions at social gatherings was not associated with an increase in R_t_.

The early estimates of R_t_ for Ghana were lower than the reported values for other countries like Nigeria,^11^ Egypt,^12^ and Kenya.^13^ The lower R values for Ghana suggests that public health and social measures were effective in containing the epidemic at the initial stage. The estimates also differ slightly from Dwomoh and colleagues (2021), but may be explained by the differences in the methods including the fact that these authors used data from the first 60 days.^14^ Nevertheless, R_t_ remained greater than 1 at the national level and in most regions indicating sustained transmission. It is therefore imperative that public health measures are strengthened throughout the country and efforts are prioritized especially in regions with larger population sizes as the disparity in the case burden across regions was reported in multiple studies.^15,16^ This disparity may be explained by the difficulty in practicing social distancing due to overcrowding stemming from high commercial activities, slum areas facilitating disease spread and urban residency. Hence, such regions will be required to implement more stringent preventive measures in order to decrease transmission.^17^

The reopening of schools was associated with a surge in cases in other jurisdictions.^18^ This finding supports the need for routine surveillance, case investigation, better protocols for isolation and quarantine, and deployment of protective personal equipment to schools. Although the correlation between changes in mobility and transmission intensity was weak in our study, decline in trips to grocery and pharmacy outlets and workplaces have been reported to be associated with lower transmission rates in most countries before and after interventions were relaxed.^19^ The difference in correlation between mobility changes and transmission intensity may be due to cultural differences, economic status, and variations in the types of interventions implemented.

It was unexpected that the relaxation of restrictions on social gatherings was not associated with increase in the R_t_ estimates, although such policies make it difficult for social distancing especially in enclosed places like churches or restaurants. This finding may be observed due to the residual effect of the prior mask mandates and the reluctance of prominent churches to resume in-person religious activities.^20^ In addition, mobility changes to retail and recreation centers remained below baseline even after the restrictions were relaxed (Figure S5).

Our study is not without limitations. First, we used publicly available data, which was subject to underreporting or reporting delays. Second, data were only available by the date of the report and unavailable by the date of symptom onset. We, therefore, accounted for this by shifting the data by nine days to approximate the time of infection. It is possible that this method of approximating the date of symptom affected the results of the cross-correlation analysis, which suggested that changes in the daily number of new infections were correlated with mobility changes 3 days later. Third, given the use of aggregate data, individual-level assumptions cannot be made. Fourth, we cannot rule out the possibility of ceiling effects during months when testing capacity was limited (Figure S6). Finally, data on socioeconomic variables was limited at the regional level hence further exploration of the case burden by region could not be performed.

In conclusion, most of the interventions implemented by the Ghanaian government were followed by an overall decrease in R_t_ estimates at the national scale, but it did not display a decline across all regions. This highlights the importance of a sustained, multi-faceted response at the national level to help mitigate the varying regional effects observed during this pandemic.

## Data Availability

All the data analyzed herein are publicly available, aggregate, data. The daily number of new infections and daily cumulative incidence data by date of report for Ghana and each of its regions are available at the Johns Hopkins University COVID-19 dashboard. The mobility data is available through Google Mobility Report.

## Co-Author Contact Information

Sylvia K. Ofori, Georgia Southern University

Jessica S. Schwind, Georgia Southern University

Kelly L. Sullivan, Georgia Southern University

Benjamin J. Cowling, The University of Hong Kong

Gerardo Chowell, Georgia State University

## Online Supplementary Materials

### Supplementary text for Methods

#### Data interpolation and moving averages

The data from obtained from the Johns Hopkins University was interpolated to estimate the values of missing data. The interpolated values were used to estimate the 3-day moving averages to reduce fluctuations in analysis.

#### Timeline for regions

At the regional level, the time series began when the first cases were reported consecutively and ended after cases were consecutively reported. For example, for Central Region, the time series began on 7^th^ May 2020. This was done to avoid estimating the *R*_*t*_ too early in the pandemic and to minimize error and uncertainty around the *R*_*t*_. Six out of the sixteen regions were included in the analysis based on having more than 4,000 cumulative cases as of December 31, 2021, to avoid the fluctuations in the estimates (see Figures S1 and S2).

#### The reproduction number, R_t_, from daily incidence using the generalized growth model (GGM)

The daily number of new infections for the first 15 days of the epidemic and selected regional levels are calibrated using the Generalized Growth Model (GGM).^1^ The GGM uses a simple generalized model for the ascending case of an epidemic using the equation:

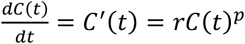

where *C*′(*t*) characterizes the epidemic curve (incident case count) over the time *t* and the solution C(t) which is the cumulative incidence at a given time *t, r* is a positive number defined as the growth rate (1/t), and *p* ∈ [0, 1] is the “deceleration of growth parameter”. At a *p* of zero, the equation describes a constant incidence and at *p*=1, the equation outlines an exponential growth dynamics. Values of *p* between 0 and 1 describe sub-exponential growth patterns.^2^

We first assume a gamma distribution for the generation interval of SARS-CoV-2 with a mean of 4.60 days and a standard deviation of 5.55 days,^3, 4^ then estimate the growth rate parameter *r*, and the deceleration of growth parameter, *p*. The GGM model simulates the growth of total cases (both imported and local cases) at *I*_*i*_, and estimates the reproduction using the renewal equation:^5^

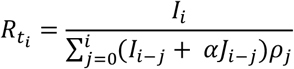

The imported cases at a given time *t* are given as *J*_*i*_; the local incidence is *I*_*i*_ at calendar time *t*_*i*_; and *ρ*_*j*_ represents the discretized probability distribution of generation interval. The factor α measures the relative contribution of imported cases to the transmission process and is assumed to be 0.15. The total number of new cases is the numerator at a given time *t= I*_*i*_, and the denominator is the total number of primary cases that generated the secondary cases. Thus, the reproduction number is the average number of new cases generated by a primary case at a given calendar time. The uncertainty around the *R*_*t*_ is derived from the parameter estimates (*r, p*). The *R*_*t*_ is estimated from 300 simulations assuming a negative binomial structure with a mean which is assumed to be a third of the variance.^2^ The *R*_*t*_ for the national curve is estimated from the first fifteen days of infection (March 12th – March 26, 2020).

#### Time-varying reproduction number *R*_*t*_, using the Instantaneous Reproduction Number method (Cori et al. method)

The *R*_*t*_ is used because it can be estimated easily in real-time and when control measures are implemented.

The instantaneous reproduction number method in the R package EpiEstim developed by Cori et al and Thompson et al. was used in the analysis.^6, 7^ The EpiEstim package uses a Bayesian framework assuming a gamma prior distribution for the posterior distribution for *R*_*t*_.^3, 6, 8^ The parametric method was used in which the serial interval was assumed to follow a gamma distribution with a mean of 4.60 days and a standard deviation of 5.55.^3, 4^

##### R_t_ using 7-day sliding windows

The instantaneous reproduction number was estimated over a 7-day sliding window given the high variation in daily *R*_*t*_ estimates. The *R*_*t*_ was assumed to be constant in each time window and the average of *R*_*t*_ estimates over 7 days was estimated with its credible intervals.

##### R_t_ using non-overlapping windows to assess the impact of various interventions

This method is used to compare the *R*_*t*_ before, during, and after a policy is implemented to assess its impact on transmission.^7^ An average was taken of the *R*_*t*_ estimates over the period between two policy change time points. The data on the major interventions and newly implemented policies were obtained from the Ghana Health Service website as well as the websites of the major news media. The following data were collected: travel bans and border closures, restriction of social activities, school closure, the mandatory wearing of masks, relaxation of restrictions on social and religious activities, deployment of personnel to monitor COVID-19 in senior high schools, reopening of air borders, and reopening of schools. A detailed list of interventions and their references can be found in Table 1. Bundled policies were implemented in close succession; hence there is difficulty in assessing the individual effects. **Regression analysis**

The power-law relationship between cumulative case count, C, and population size, N (i.e., C∼N^g^ where g is an exponent) was transformed into a relationship between log_10_-transformed per-capita cumulative case count and log_10_-transformed population size as follows:^9^

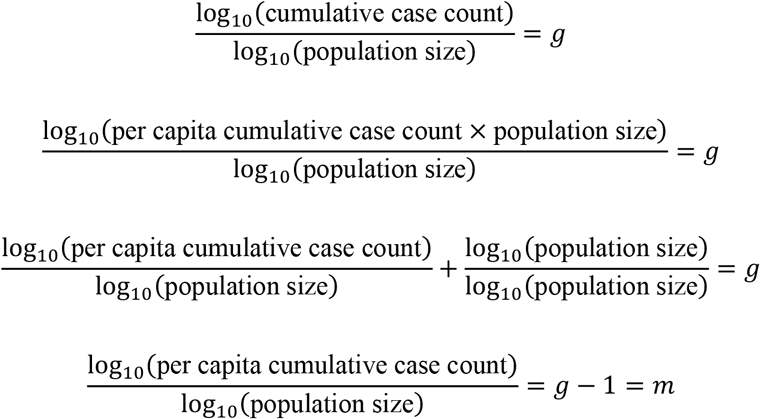

The per capita cumulative case count was calculated by dividing the total cumulative case count by the population size of the nation or region at the specified time point. A linear regression model was fit to the data of log_10_-transformed per capita cumulative case count and log_10_-transformed population size. The slope *m* of the regression line (where *m*=*g*-1) can be interpreted as follows: if *m*=0, there is a homogeneity of per capita cumulative case count across regions; if *m*<0, low-population regions have higher per capita cumulative case count than high-population regions; and if *m*>0, low-population regions have lower per capita cumulative case count than high-population regions.^9^ The 2020 population estimates for Ghana and the regions were obtained from the Ghana Statistical Service.^10^

#### Google mobility data

The mobility data was sourced from Google to analyze changes in the number of visits to places in the following categories: (a) grocery and pharmacy, (b) parks, (c) transit stations, (d) retail and recreation, (e) residential, and (f) workplaces.^11^ The data provides information for how visits and duration of staying at different places changes compared to baseline values which is the median value for the corresponding day of the week, during the five weeks of January 3 — February 6, 2020. The data is representative of users who opted for location history. The relationship between the 7-day moving average of mobility changes in retails and recreation, grocery and pharmacy, and workplace, and the 3-day moving average of the daily number of new infections (or *R*_*t*_ estimates from a 7-day sliding window) is assessed using the time-lagged cross-correlation using the assumed dates of infection.

## Supplementary text for Results

### Epidemic curves

Four waves were observed in the epidemic curves for Ghana. The daily number of new cases of COVID-19 in Ghana, the Greater Accra Region, and the Ashanti region decreased from mid-May to mid-June and surged in mid-July, 2020. For all three locations, the daily number of new cases dropped again to less than 500 per day and spiked again in January 2021 (Figure 1). Relatively few new cases were observed between March and July 2021, before rising again in late July. Very high numbers of new cases were recorded from December 2021 due to Christmas festivities. The Western region’s daily new cases were highest in June and July 2020, and January-February 2021 (Figure S3). The daily number of new cases for the Central, Volta, and Eastern regions fluctuated throughout the study period but followed a similar trajectory with that of Ghana as a whole in the peak periods (Figures S3).

### Comparing R_t_ Estimates using GGM and Cori et al. method

Using the GGM, the COVID-19 *R*_*t*_ estimate for Ghana was estimated at 1.8 (95% CI: 1.7, 2) for the first 15 days of the epidemic (Figure S4) compared to that estimated using the Cori et al. method, which estimated the *R*_*t*_ at 1.85 (95% CI: 1.42, 2.27).

The difference in estimates may be because in the Cori et al. method, fluctuations in daily number of new infections are treated as true reflections of the underlying epidemic while the GGM method treats these fluctuations as noise by assuming an error structure underneath. Hence, the estimated value of *R*_*t*_ using the Cori et al. method may provide a better reflection of how early interventions and policy changes affected the *R*_*t*_ at the national level for this study.

---

**Table S1:** Cumulative number of cases and cumulative incidence rate by region as of December 31, 2021 (N= 142,986)

| **Region** | **Population** | **Cumulative Number of Cases** | **Cumulative Number of Cases per 100,000** |
| --- | --- | --- | --- |
| Ghana (national) | 32,000,000 | 142,986 | 446.83 |
| Ahafo | 613,000 | 1,074 | 175.20 |
| Ashanti | 5,924,500 | 21,646 | 365.36 |
| Bono | 1,168,800 | 2,119 | 181.30 |
| Bono East | 1,133,800 | 2,667 | 235.23 |
| Central | 2,605,500 | 5,143 | 197.39 |
| Eastern | 3,318,900 | 6,661 | 200.70 |
| Greater Accra | 5,055,900 | 79,567 | 1573.75 |
| North East | 588,800 | 283 | 48.06 |
| Northern | 1,948,900 | 1,787 | 91.69 |
| Oti | 759,800 | 850 | 111.87 |
| Savannah | 594,700 | 263 | 44.22 |
| Upper East | 1,302,700 | 1,484 | 113.92 |
| Upper West | 868,500 | 1,543 | 177.66 |
| Volta | 1,907,700 | 5,621 | 294.65 |
| Western | 2,214,700 | 7,817 | 352.96 |
| Western North | 949,100 | 1,049 | 110.53 |

**Table S2:** Median estimates and 95% credible intervals of nonoverlapping window *R*_*t*_ for each period between each policy change (left) and percent changes in nonoverlapping window *R*_*t*_ and associated 95% credible intervals with given policies.

| | $R_t$ | | Percentage change (%) | |
| --- | --- | --- | --- | --- |
| Policy change | Median | 95% Credible Interval | Median | 95% Credible Interval |
| **National** |  |  |  |  |
| Before A | 1.92 | 1.54, 2.34 |  |  |
| A to B | 1.54 | 1.42, 1.67 | -20.19 | -34.75, +3.91 |
| B to L | 1.05 | 0.94, 1.16 | -32.63 | -41.19, -22.87 |
| L to C | 1.31 | 1.29, 1.34 | +25.62 | +13.60, +40.42 |
| C to D | 1.05 | 1.04, 1.06 | -19.97 | -21.81, -18.05 |
| D to E | 1.09 | 1.09, 1.10 | +4.03 | +2.61, +5.49 |
| E to F | 0.92 | 0.91, 0.93 | -15.95 | -16.86, -15.10 |
| F to G | 1.04 | 1.03, 1.05 | +13.01 | +11.74, +14.25 |
| G to V | 1.03 | 1.03, 1.04 | -0.60 | -1.58, 0.49 |
| V to H | 0.98 | 0.98, 0.99 | -4.80 | -5.51, -4.09 |
| Beyond H | 1.09 | 1.08, 1.10 | +10.57 | +9.62, +11.47 |
| **Regions** |  |  |  |  |
| **Greater Accra** |  |  |  |  |
| Before A | 1.75 | 1.35, 2.21 |  |  |
| A to B | 1.62 | 1.47, 1.77 | -7.48 | -27.84, +20.48 |
| B to L | 1.17 | 1.06, 1.29 | -27.41 | -36.26, -16.68 |
| L to C | 1.27 | 1.23, 1.29 | +7.70 | -2.70, +19.31 |
| C to D | 1.00 | 0.99, 1.02 | -20.84 | -22.88, -18.75 |
| D to E | 1.06 | 1.05, 1.07 | +6.02 | +4.05, +7.78 |
| E to F | 0.95 | 0.94, 0.96 | -10.07 | -11.45, -8.68 |
| F to G | 1.03 | 1.02, 1.04 | +8.24 | +6.87, +9.64 |
| G to V | 0.98 | 0.97, 0.99 | -5.30 | -6.47, -4.30 |
| V to H | 1.01 | 1.00, 1.02 | +3.43 | +2.35, +4.46 |
| Beyond H | 1.47 | 1.45, 1.49 | +45.15 | +43.33, +46.99 |
| **Ashanti** |  |  |  |  |
| Before A | 1.84 | 1.20, 2.69 |  |  |
| A to B | 0.61 | 0.29, 1.11 | -66.67 | -85.65, -32.54 |
| B to L | 2.15 | 1.50, 2.96 | +250.86 | +84.23, +681.67 |
| L to C | 1.40 | 1.27, 1.54 | -35.34 | -52.66, -6.37 |
| C to D | 1.13 | 1.10, 1.16 | -19.34 | -26.19, -11.69 |
| D to E | 1.03 | 1.02, 1.05 | -8.49 | -11.08, -5.82 |
| E to F | 0.95 | 0.93, 0.96 | -8.43 | -10.45, -6.53 |
| F to G | 1.00 | 0.95, 1.05 | +5.75 | +0.35, +10.98 |
| G to V | 1.03 | 1.01, 1.05 | +2.87 | -2.34, +8.52 |
| V to H | 0.98 | 0.97, 0.99 | -4.84 | -6.67, -2.27 |
| Beyond H | 1.45 | 1.39, 1.50 | +47.31 | +41.77, +52.84 |
| **Central** |  |  |  |  |
| Before C | 1.47 | 1.04, 1.99 |  |  |
| C to D | 1.16 | 1.10, 1.22 | -20.86 | -42.14, +14.15 |
| D to E | 1.06 | 1.00, 1.09 | -9.65 | -15.07, -3.46 |
| E to F | 0.94 | 0.91, 0.97 | -9.99 | -14.28, -5.25 |
| F to G | 0.94 | 0.88, 0.99 | -0.58 | -7.17, +6.52 |
| G to V | 1.16 | 1.12, 1.20 | +23.31 | +15.66, +32.75 |
| V to H | 0.95 | 0.93, 0.97 | -17.86 | -21.02, -14.39 |
| Beyond H | 1.24 | 1.00, 1.51 | +29.70 | +5.24, +59.34 |
| **Eastern** |  |  |  |  |
| Before C | 1.21 | 1.09, 1.34 |  |  |
| C to D | 1.21 | 1.11, 1.32 | +0.63 | -12.97, +15.60 |
| D to E | 1.05 | 1.01, 1.08 | -13.89 | -21.62, -4.77 |
| E to F | 0.99 | 0.96, 1.01 | -5.54 | -9.76, -1.18 |
| F to G | 0.88 | 0.84, 0.94 | -10.15 | -15.58, -4.52 |
| G to V | 1.05 | 1.02, 1.07 | +17.86 | +10.66, +26.23 |
| V to H | 0.98 | 0.96, 0.99 | -6.73 | -9.74, -3.58 |
| Beyond H | 1.55 | 1.35, 1.76 | +58.30 | +40.16, +80.74 |
| **Volta** |  |  |  |  |
| Before C | 1.73 | 1.40, 2.12 |  |  |
| C to D | 1.43 | 1.33, 1.54 | -18.29 | -31.95, +3.86 |
| D to E | 0.85 | 0.80, 0.91 | -40.35 | -45.64, -34.66 |
| E to F | 0.90 | 0.84, 0.96 | +4.92 | -4.71, +15.57 |
| F to G | 1.06 | 0.97, 1.16 | +18.83 | +5.92, +31.69 |
| G to V | 1.08 | 1.04, 1.12 | +1.06 | -7.43, +11.28 |
| V to H | 0.97 | 0.95, 0.99 | -10.08 | -12.68, -6.92 |
| Beyond H | 2.20 | 2.02, 2.39 | +127.92 | +109.66, +148.11 |
| **Western** |  |  |  |  |
| Before C | 1.53 | 1.24, 1.87 |  |  |
| C to D | 1.26 | 1.22, 1.30 | -18.77 | -33.71, +0.40 |
| D to E | 0.97 | 0.94, 0.99 | -23.26 | -26.55, -19.97 |
| E to F | 0.85 | 0.82, 0.88 | -11.71 | -15.88, -7.83 |
| F to G | 1.12 | 1.05, 1.19 | +31.58 | +22.22, +41.45 |
| G to V | 1.04 | 1.02, 1.06 | -7.49 | -13.28, -1.24 |
| V to H | 0.95 | 0.93, 0.97 | -8.37 | -11.05, -5.64 |
| Beyond H | 1.91 | 1.79, 2.03 | +101.03 | +88.67, +114.64 |

**Table S3:** Linear regression analysis between the log_10_-transformed per capita cumulative case count and log_10_-transformed population size using cumulative incidence for sixteen regions.

| **Time point** | **Slope** | **95% confidence interval** | **P value** |
| --- | --- | --- | --- |
| June 30, 2020 | 1.59 | 0.91, 2.28 | 0.0002 |
| August 31, 2020 | 1.17 | 0.53, 1.82 | 0.0016 |
| October 31, 2020 | 1.19 | 0.54, 1.84 | 0.0014 |
| December 31, 2020 | 1.21 | 0.57, 1.85 | 0.0011 |
| February 28, 2021 | 1.10 | 0.62, 1.58 | 0.000247 |
| April 30, 2021 | 0.94 | 0.53, 1.35 | 0.000216 |
| June 30 2021 | 0.94 | 0.52, 1.35 | 0.00024 |
| August 31 2021 | 0.91 | 0.48, 1.34 | 0.00050 |
| October 31 2021 | 0.86 | 0.45, 1.28 | 0.00052 |
| December 31 2021 | 0.90 | 0.47, 1.33 | 0.00048 |

**Table S4:** Time-lag correlation coefficients between the 3-day moving average of daily number of new COVID-19 cases, and 7-day moving average of relative mobility (percentage change from baseline) for trips to retails and recreation, grocery and pharmacy, workplace, and residential categories over the first 90 days of the pandemic in Ghana using the assumed date of infection.^b^

| Lag (days) | Retails and recreation |  | Grocery and pharmacy |  | Workplace |  | Residential |  |
| --- | --- | --- | --- | --- | --- | --- | --- | --- |
|  | r | P value | r | P value | r | P value | r | P value |
| -3 | 0.042 | 0.693 | 0.172 | 0.103 | 0.118 | 0.263 | 0.067<sup>a</sup> | 0.528 |
| -2 | 0.069 | 0.516 | 0.205 | 0.052 | 0.149 | 0.159 | 0.044 | 0.629 |
| -1 | 0.096 | 0.363 | 0.237 | 0.024 | 0.178 | 0.091 | 0.022 | 0.836 |
| 0 | 0.123 | 0.242 | 0.271 | 0.010* | 0.209 | 0.047* | 0 | 0.999 |
| 1 | 0.167 | 0.113 | 0.307 | 0.004* | 0.248 | 0.019* | -0.042 | 0.687 |
| 2 | 0.208 | 0.048* | 0.341 | 0.001* | 0.291 | 0.006* | -0.085 | 0.418 |
| 3 | 0.243<sup>a</sup> | 0.021* | 0.366<sup>a</sup> | 0.001* | 0.328<sup>a</sup> | 0.002* | -0.124 | 0.240 |

\*p<0.05
<sup>a</sup> The model with the largest (absolute value) correlation coefficient in each category.
<sup>b</sup> Data for the first 90 days was used to observe the impact of relaxation of social restrictions on mobility changes.

**Table S5:** Time-lag correlation coefficients between median 7-day sliding window *R*_*t*_ of COVID-19 and 7-day moving average of relative mobility (percentage change from baseline) for trips to retails and recreation, grocery and pharmacy, workplace, and residential categories over the first 90 days of the pandemic in Ghana using the assumed date of infection.^b^

| **Lag**<br>**(days)** | **Retails and**<br>**recreation** |  | **Grocery and**<br>**pharmacy** |  | **Workplace** |  | **Residential** |  |
| --- | --- | --- | --- | --- | --- | --- | --- | --- |
|  | **r** | **P value** | **r** | **P value** | **r** | **P value** | **r** | **P value** |
| **-3** | 0.204<sup>a</sup> | 0.053 | 0.061<sup>a</sup> | 0.562 | 0.137 | 0.192 | -0.295 | 0.005* |
| **-2** | 0.203 | 0.054 | 0.055 | 0.602 | 0.139<sup>a</sup> | 0.188 | -0.310 | 0.003* |
| **-1** | 0.197 | 0.062 | 0.042 | 0.691 | 0.130 | 0.216 | -0.317 | 0.003* |
| **0** | 0.197 | 0.061 | 0.031 | 0.766 | 0.124 | 0.240 | -0.328<sup>a</sup> | 0.002* |
| **1** | 0.164 | 0.119 | 0.004 | 0.972 | 0.099 | 0.346 | -0.297 | 0.005* |
| **2** | 0.133 | 0.207 | -0.020 | 0.851 | 0.072 | 0.497 | -0.264 | 0.012* |
| **3** | 0.108 | 0.304 | -0.033 | 0.755 | 0.051 | 0.631 | -0.238 | 0.024* |

\* $p < 0.05$ <sup>a</sup> The model with the largest (absolute value) correlation coefficient in each category.
<sup>b</sup> Data for the first 90 days was used to observe the impact of relaxation of social restrictions on mobility changes.

## Supplemental Figure Captions and Legends

**Figure S1:**
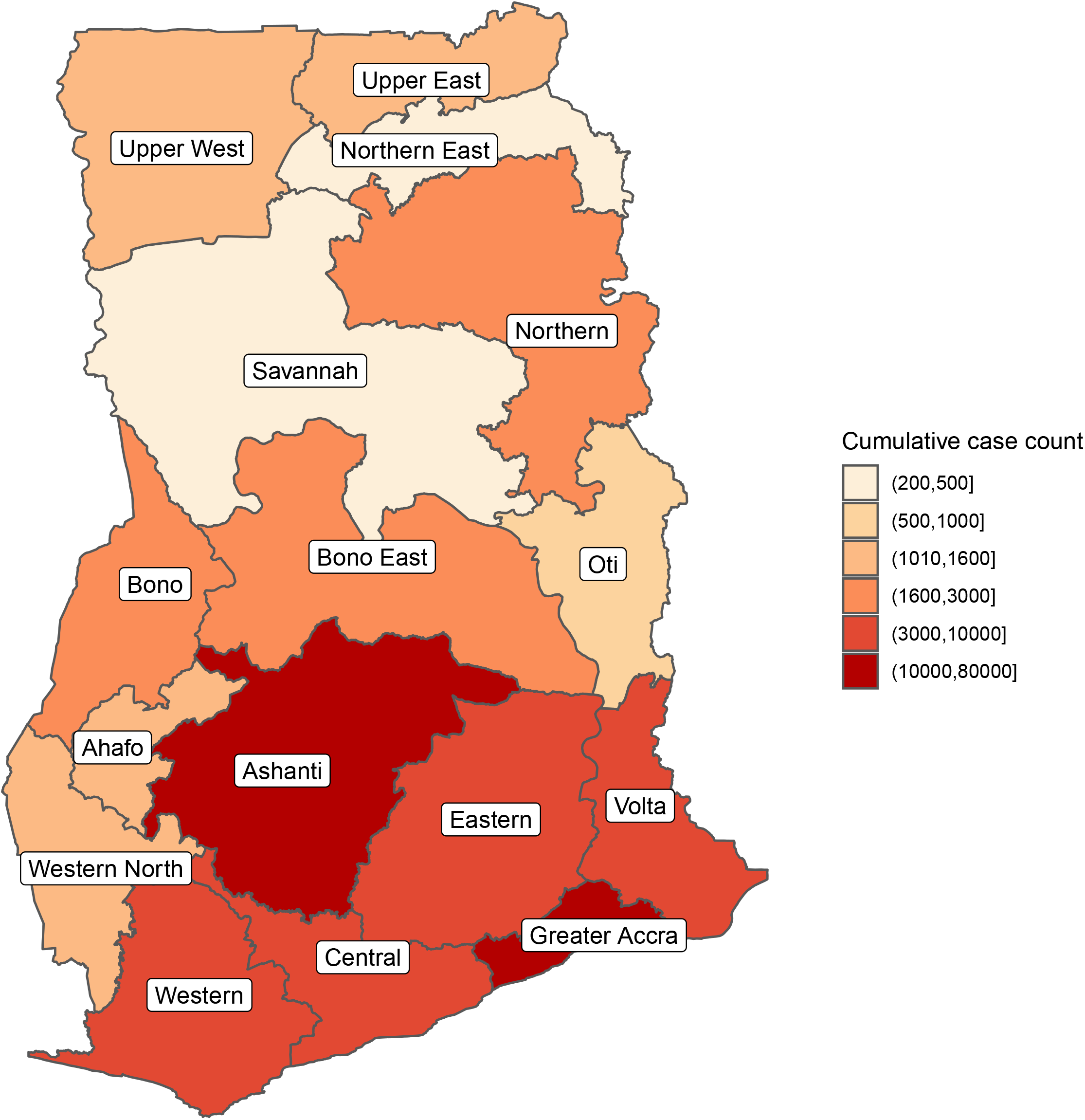
Cumulative case count as of December 31, 2021 in Ghana by region.

**Figure S2:**
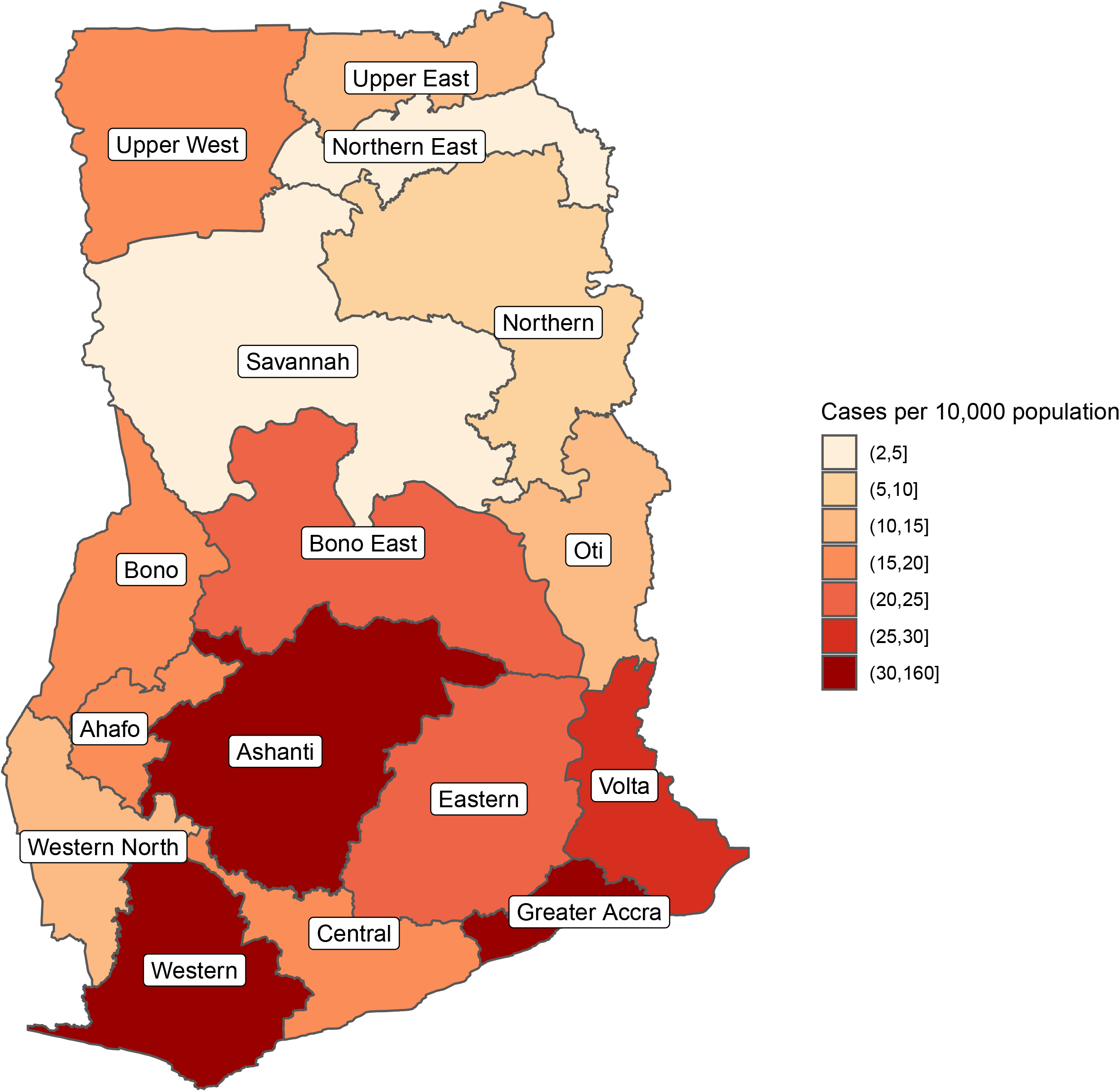
Cumulative case count per 10,000 population as of December 31, 2021 in Ghana by region.

**Figure S3:**
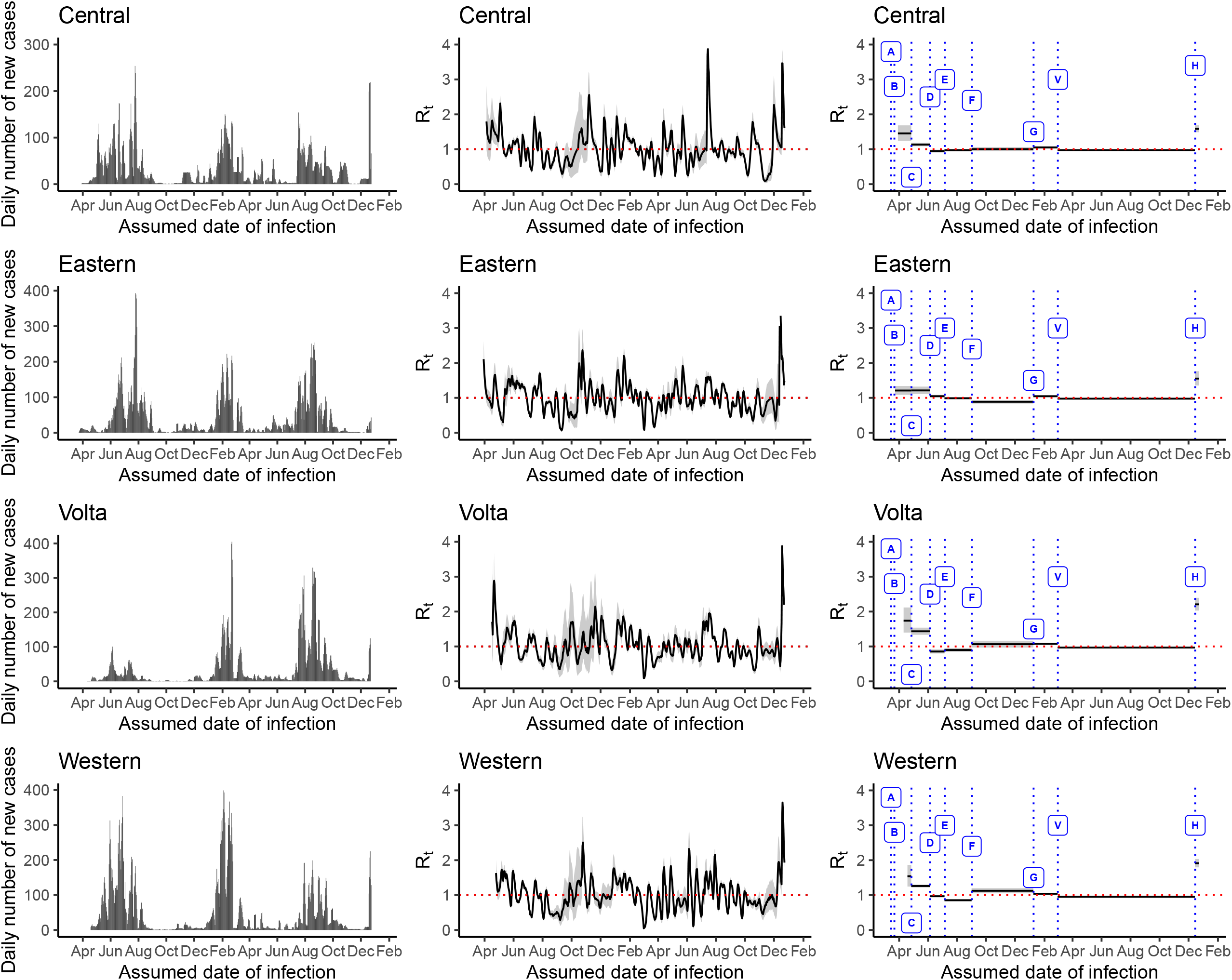
The daily number of new cases (left panel), 7-day sliding window *R*_*t*_ (middle panel) and nonoverlapping window *R*_*t*_ (right panel) estimated using the Cori et al. method in the ‘EpiEstim’ package, in the Central, Eastern, Volta and Western regions, March 12, 2020— December 31, 2021. The government policies represented by the alphabets in the figure are: A = restrictions of all travels to Ghana, suspension of social gatherings, school closure, mandatory 14-day quarantine for all travelers; B = closure of all borders to human traffics; C = mandatory wearing of facemasks at all businesses and organizations; D = relaxation of restrictions at social gatherings; E = deployment of personnel to monitor COVID-19 cases in high schools; F = reopening of international borders; G= reopening of schools, V = vaccination rollout, H = Christmas festivities in 2021.

**Figure S4:**
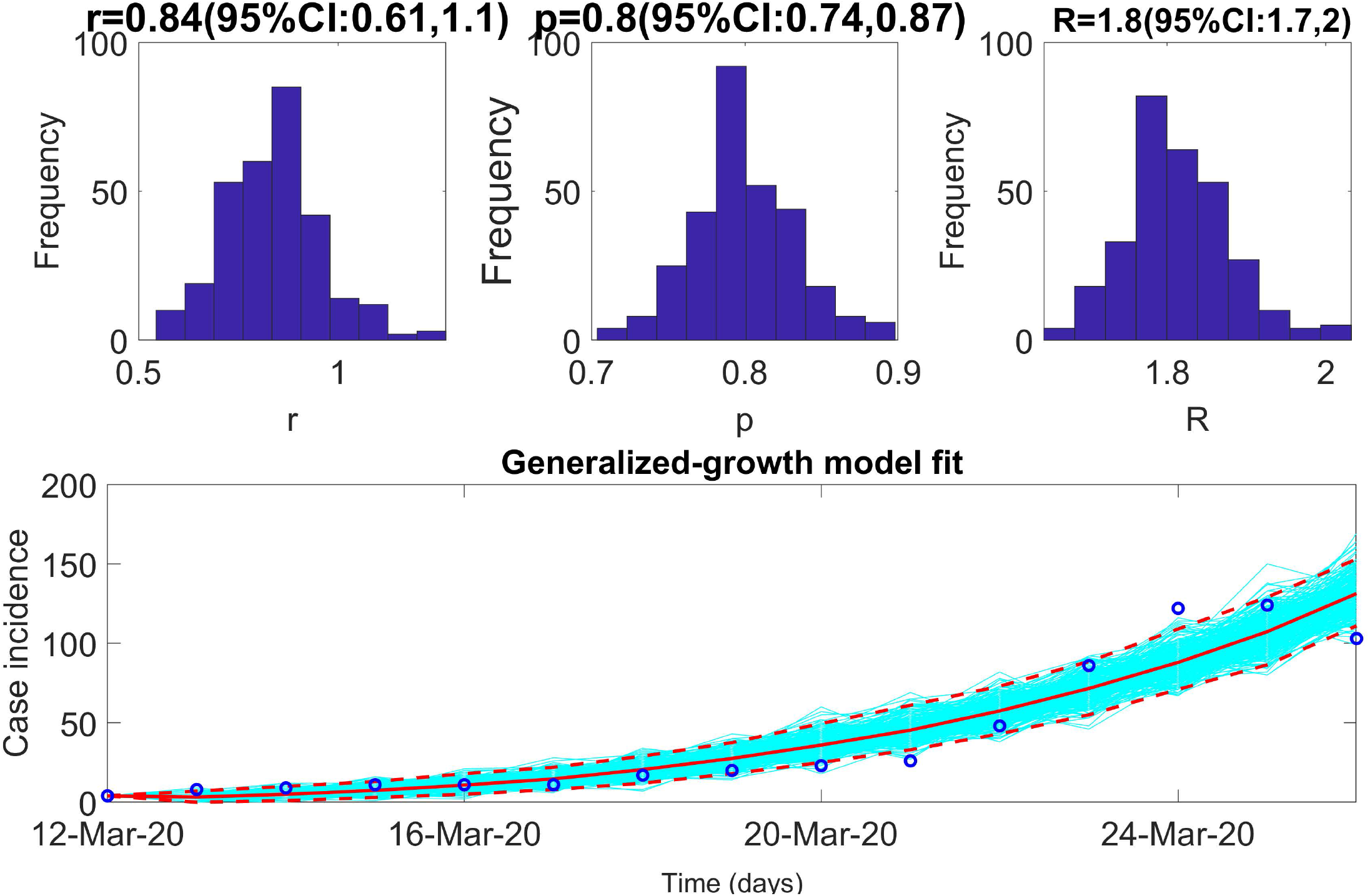
Results from the Generalized Growth Model. Upper panel shows the estimated reproduction number of 1.8 (95% CI: 1.7, 2) for Ghana with 95% CI from March 12, 2020, to March 26, 2020. The growth rate, *r*, 0.84 (95% CI: 0.61, 1.1), deceleration growth parameter, *p*, 0.8 (95% CI: 0.74, 0.87). The lower panel shows the GGM fit of the daily reported number of new infections using the first 15 days of data.

**Figure S5:**
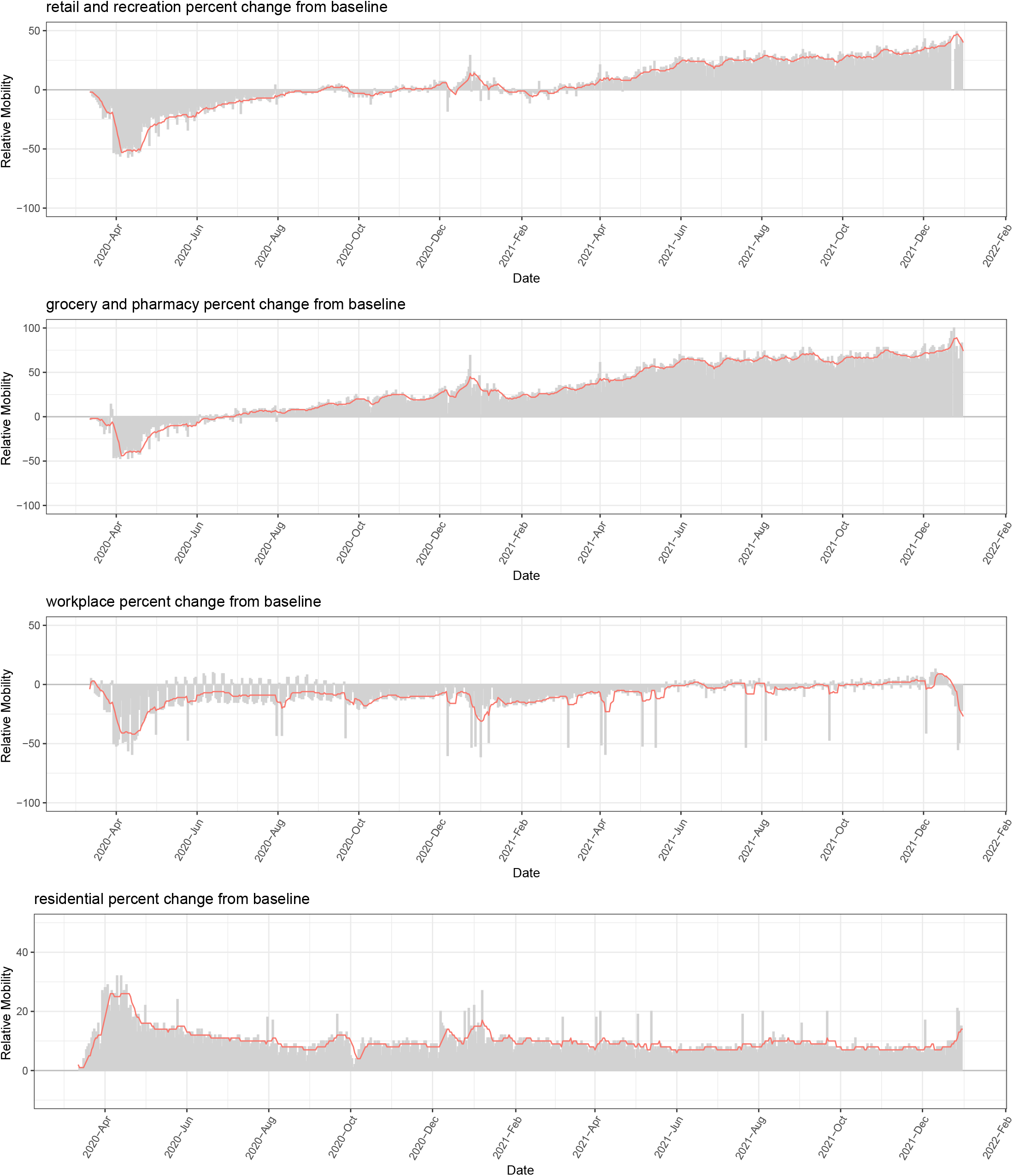
Google mobility trends by type of mobility: retail and recreation (first panel), grocery and pharmacy (second panel), workplaces (third panel) and residential (fourth panel) from February 15, 2020, to December 31, 2021. Both the original data (bar) and the 7-day moving average of mobility data (red line) are displayed.

**Figure S6:**
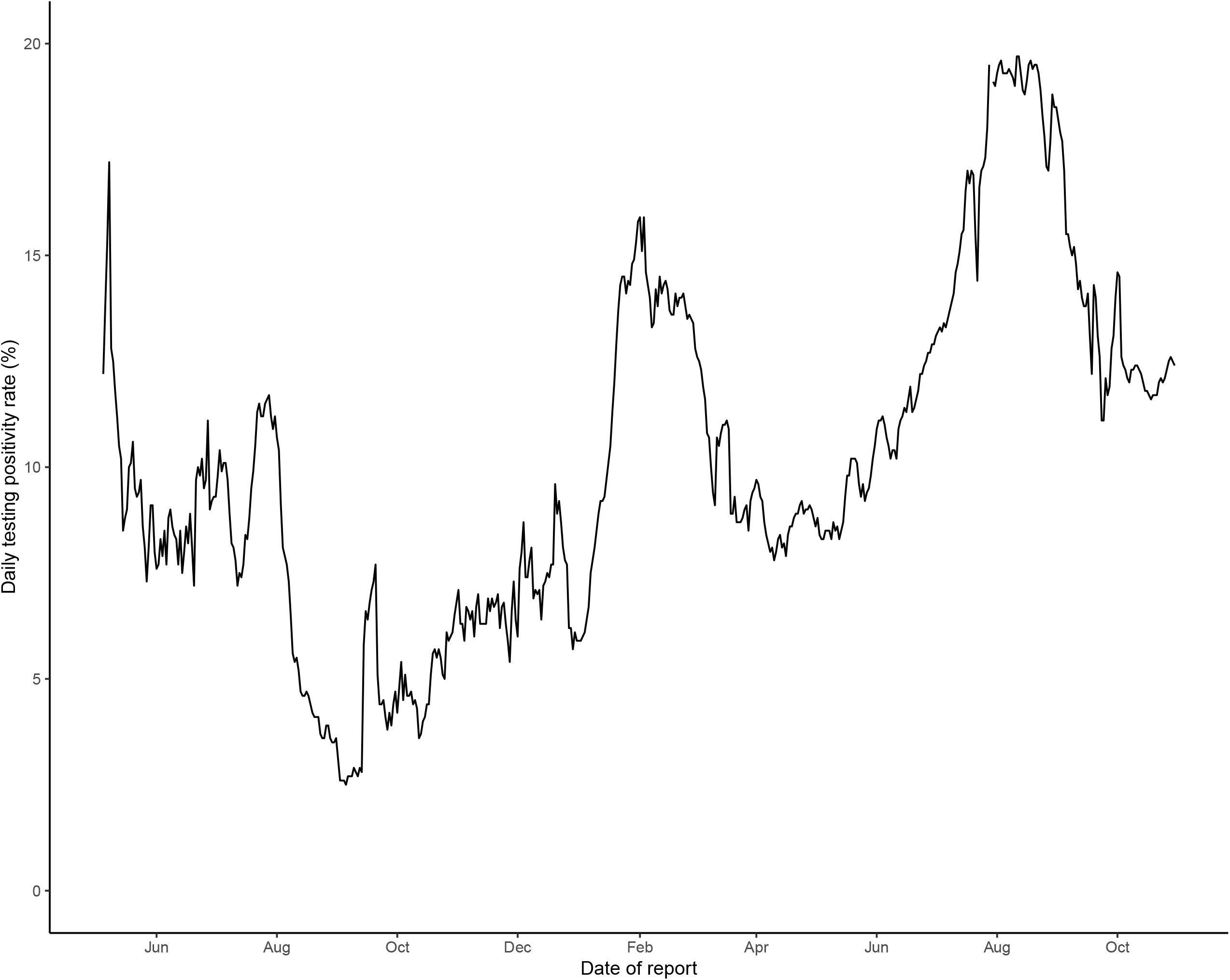
COVID-19 testing positivity rate in Ghana by date of report from May 5, 2020 to October 30, 2021.

